# Prioritizing protein targets for dyslipidaemia and cardiovascular diseases using Mendelian randomization in South Asians

**DOI:** 10.1101/2024.12.18.24319223

**Authors:** Siwei Wu, Alexander Smith, Jingxian Huang, Georg W. Otto, Yi-Hsuan Ko, James Yarmolinsky, Dipender Gill, Anand Rohatgi, Abbas Dehghan, Ioanna Tzoulaki, Devendra Meena

## Abstract

South Asians are at higher risk of dyslipidaemia–a modifiable risk factor for cardiovascular diseases (CVDs). We aimed to identify protein targets for dyslipidaemia and CVDs in this population.

We used a two-sample Mendelian randomization (MR) approach, supplemented with MR-Egger, weighted median, colocalization, and generalized MR (GMR), to evaluate the effect of 2,800 plasma proteins on high/low/non-high-density lipoprotein cholesterol (HDL-C/LDL-C/nonHDL-C), total cholesterol, and triglycerides. Observational analyses were conducted on MR findings with strong colocalization (posterior probability≥ 80%) and GMR findings. Univariate MR assessed lipid-associated proteins’ effect on CVDs. Finally, we compared the potential causal effects of plasma proteins on lipids in South Asians with those in Europeans to study heterogeneity in the MR effects.

We identified 29 genetically proxied proteins potentially causal to at least one lipid measure, 12 of which showed strong colocalization and GMR evidence, including ANGPTL3 and PCSK9. Notably, PCSK9 demonstrated a stronger association with LDL-C in European compared to South Asian (β_European_= 0.37; 95% Confidence Interval (CI)= (0.36, 0.38), β_South Asian_= 0.16; 95% CI= (0.11, 0.21)). Observational analysis suggested significant interaction between PCSK9 levels with LDL-C levels in South Asians with South Asians having a significantly lower effect compared to other ethnicities (PCSK9*South Asian; β= -0.14; 95% CI= (-0.174, -0.107)). Additionally, we showed that CELSR2 is also linked with CAD in South Asians. Our study highlighted potential causal links between plasma proteins, dyslipidaemia, and CVD in South Asians, with significant heterogeneity across genetic ancestry groups. Larger studies in South Asians are needed to validate these findings.

## Introduction

Routinely measured lipid parameters including high/low/non-high-density lipoprotein cholesterol (HDL-C/LDL-C/nonHDL-C), total cholesterol (TC), and triglycerides (TG), are well-established risk factors for cardiovascular diseases (CVDs), including coronary artery disease (CAD), stroke, and heart failure, (1–4) all of which are leading causes of mortality and morbidity worldwide (5). HDL-C and TC have been included in several risk equations like SCORE and PCE as important predictors for CVDs (6, 7). The role of HDL-C and TG as the target for CVD intervention is inconclusive but under investigation (8, 9). LDL-C and nonHDL-C are not only prognostic risk markers but also established therapeutic targets for CVDs (10, 11). Building on this, evidence suggests that individuals of South Asian ancestry, compared with non-Hispanic white population, have a higher prevalence of dyslipidaemia (12) and are more susceptible to cardiometabolic diseases closely related to dyslipidaemia including CAD (13, 14), stroke (15), and type 2 diabetes (16). Therefore, understanding the genomic and proteomic makeup of plasma lipids and identifying causal factors in South Asians is crucial for intervening in dyslipidaemia and preventing lipid-related cardiovascular conditions although the residual risk of lipid modifying medications remains.

Circulating plasma proteins are key to disease mechanisms and are promising drug targets. Previous studies have identified proteins associated with lipids some of which are targeted for dyslipidaemia treatment (17). The most prominent example is Apolipoprotein B (APOB), which is one of the most prognostic and best therapeutic targets for CVDs (18). Despite recent advances, most plasma proteins linked to lipids have been discovered in the European population (19). The proteomic findings in non-European individuals are limited, especially in South Asians, which is the most rapidly growing but neglected population with higher susceptibility to dyslipidaemia (12). We recently reported that certain plasma proteins may exert ancestry-specific causal effects on certain CVDs (20). Therefore, understanding the proteomic features of dyslipidaemia in South Asians is essential for developing more effective disease prevention strategies and drug discovery approaches for this high-risk population.

Recent genome wide association studies (GWAS) involving South Asian populations have made resources on circulating plasma proteins and lipid traits available (21, 22). Furthermore, advances in methodologies for causal inference in epidemiology, including the Mendelian randomization (MR) framework, now allow us to investigate the potential causal relationship between an exposure (e.g. plasma proteins) and an outcome (e.g. lipid traits) for which GWAS summary statistics are available (23). MR utilises genetic variants as instrument variables (IVs), and is less susceptible to confounders and reverse causation bias than other study designs, and can be applied to for causal inference (24). Such approach was applied to systematically evaluate plasma proteins’ effect on lipid traits in EUR, but not yet in SAS (25).

In this study, we aimed to (1) systematically evaluate bi-directional causal effects of plasma proteins on five lipid traits in South Asians using a two-sample proteome-wide MR approach (2) Investigate whether lipid-associated plasma proteins affect CAD and stroke risk in South Asians (3) Compare the effects of plasma proteins on lipid fractions in South Asians and Europeans.

## Method

### Data source

#### Genetic associations for genetically predicted plasma proteins

In the UK Biobank Pharma Proteomics Project (UKBPPP), 2,940 probes capturing 2,922 unique proteins were made available (22). We defined each probe as a protein and extracted genetic associations of the 2,940 plasma proteins in individuals of Central/South Asian ancestry (CSA, N=920) and European ancestry (EUR, N=34,557) from the UKBPPP (22). The ancestries were defined with the pan-UK Biobank (UKBB) definitions of genetic ancestry (available in UKBB return dataset 2442). We included the 2,800 plasma proteins as the primary exposures after excluding the ones whose cognate gene is ambiguous (N = 15) and those encoded by genes lying on the X chromosome (due to unavailability of X chromosome data on outcome; N = 88) or the MHC region (due to complex LD in this region; N = 37).

### Genetic associations for genetically predicted lipid traits and CVDs

Five lipid traits including HDL-C, LDL-C, nonHDL-C, TC, and TG were included as the primary outcome in MR analysis. Ancestry-specific (N_SAS_C=Cup to 40,963, N_EUR_C=C1,320,016) genetic associations of the 5 traits were publicly available from the Global Lipids Genetics Consortium (21). The GWAS statistics for CAD were obtained from the East London Genes & Health (ELGH) study for SAS (26) and from the CARDIoGRAMplusC4D Consortium for EUR (27). The GWAS summary data on stroke for EUR and SAS were sourced from the GIGASTROKE consortium (28).

### Proteome-wide MR and colocalization analysis on lipid fractions in SAS Instrument selection and MR

To obtain genetic instruments for plasma proteins, biallelic single nucleotide polymorphisms (SNP) lying within +/- 500 kilobase (KB) from the coding gene (defined as cis-acting SNPs), with minor allele frequency (MAF) > 0.05, and reaching genome wide significant level (P < 5 × 10^-8^) were extracted. The SNPs were further harmonized to the 5 lipid fractions and clumped to an r^2^ < 0.001 to ensure independence across the instruments. Subsequently, a F- statistic was calculated for each SNP and SNPs with F-statistic < 10 were excluded to avoid weak instrument bias (29). We also applied the Steiger filtering and excluded the SNPs with potential reverse causality (30). After applying these filters, we ended up with 708 proteins with at least 1 instrument available which were carried forward for the downstream analysis (Figure S1). The Wald ratio method was applied to proteins with 1 single SNP as the instrument while the inverse-variance weighted (IVW) model was applied to proteins which were instrumented by 2 or more SNPs (23). To test for horizontal pleiotropy and to ensure robustness of the proteome-wide MR findings, we applied MR-Egger and Weighted median to the significant associations where at least 3 instruments were available (31, 32). For multiple testing correction, a false discovery rate (FDR) was calculated by applying the Benjamini-Hochberg (BH) adjustment to each lipid fraction (33) and an FDR < 0.05 was defined to be significant.

To avoid bias due to sample overlap between UKBPPP and GLGC, we also performed MR using GLGC data excluding UKBB participants. A correlation analysis was performed to compare the beta estimates derived from GLGC data with or without UKBB individuals.

### Bayesian colocalization

A Bayesian colocalization analysis was conducted on all protein-lipid associations with FDR-corrected P < 0.05 to determine if they shared the same causal variant (34). The colocalization enabled us to minimise horizontal pleiotropy caused by linkage disequilibrium (LD) where the plasma protein levels and lipid traits were influenced by 2 distinct variants in LD with each other (35). Colocalization was performed on the same window as the previous proteome-wide MR (within +/- 500KB of the cognate gene), with rare variants (MAF < 0.05) residing in the window dropped. Default priors as described in the original paper were applied (34). A posterior probability of colocalization (PPH4) ≥ 80% indicated strong colocalization, while 60% ≤ PPH4 < 80% was considered suggestive evidence for colocalization.

### MR generalized to correlated instruments (GMR)

Due to stringent genetic instrument selection, most proteins had few pQTLs, which could bias the MR estimates due to unknown pleiotropy and make MR-Egger and WM unapplicable. Therefore, to improve robustness, we included additional instruments at p < 1 × 10CC and clumped them with r² = 0.4 (36). To account for correlation between instruments, a generalized inverse variance weighted regression (gIVW) was applied as the primary method (37). Where applicable, we also performed the MR-Egger generalized to correlated variants (gEgger) (38) and weighted median (32).

### Causal effects of lipid-associated proteins on CAD and stroke

A *cis*-MR analysis was conducted with lipid-associated plasma proteins (P-FDR< 0.05) as exposures and CAD and stroke (any stroke, any ischemic stroke, large artery stroke, cardioembolic stroke, and small vessel stroke) as outcomes. An FDR correction was applied separately for each outcome while colocalization and supplemental MR methods (gIVW, gEgger, and weighted median) were performed to validate the genetically proxied associations surviving the FDR correction (32–34, 37, 38). The instrument selection criteria and parameters for MR and colocalization in this step were set as described above.

Additionally, univariate MR was performed to assess whether the lipids measures show similar causal effects on CVDs in South Asians compare to Europeans. Genetic instruments for lipid fractions were extracted from the whole genome (autosomes only), and other MR criteria and methods were the same as those applied in *cis*-MR (29, 30). Where applicable, MR-Egger and weighted median were applied as well (31, 32).

Furthermore, for plasma proteins associated with both lipids and CVDs, we evaluated with multi-trait colocalization whether the 3 traits have a shared causal variant in the corresponding gene region (39). The colocalization analysis was applied on plasma protein, lipid fractions, and the CVD on the genomic region +/- 500KB extended from the cognate gene (39). Rare variants with MAF < 0.05 were dropped. Priors for multi-trait colocalization were set to default values: the probability of any SNP within the colocalization window being exclusively associated with one of the three traits was 1 × 10CC, with two traits was 1 × 10CC, and with all three traits was 1 × 10CC(39).

### Reverse MR with lipid fractions as exposures and plasma proteins as outcomes in SAS

Furthermore, to understand the potential causal effects of dyslipidaemia on plasma protein abundance, we conducted reverse MR using lipid fractions (exposure) and plasma proteins (outcomes). Genetic instruments were selected from the 22 autosomes using the filtering criteria as previously described.

### Comparison with European population

For plasma proteins with potential effects on lipid fractions in SAS, we estimate their effects in EUR using a two-sample MR approach, applying the same genetic instrument criteria used in SAS. We checked the consistency of the MR estimates and applied a correlation analysis between EUR and SAS population estimates for proteins that were consistent and significant in both groups. We also compared the 95% confidence intervals, defining a significant difference in the genetically proxied MR estimates when the intervals did not overlap.

### Observational associations of protein levels with lipid fractions

For plasma proteins associated to LDL-C and HDL-C with all the MR, strong colocalization, and GMR evidence, we conducted observational analysis using linear regression for each protein and its associated lipid fraction. Each regression model was adjusted for age, sex, Townsend deprivation, BMI, HbA1c, cholesterol medication, smoking, systolic blood pressure, blood pressure medication, and ethnicity. To evaluate potential effect modification by ancestry, we included an interaction term between a binary variable for South Asian ancestry and protein level. All continuous variables and lipid fractions were standardised before modelling and p values were adjusted for multiple testing using FDR at 5%.

## Result

An overview of the study design is shown in **Figure 1**.

**Figure 1.**
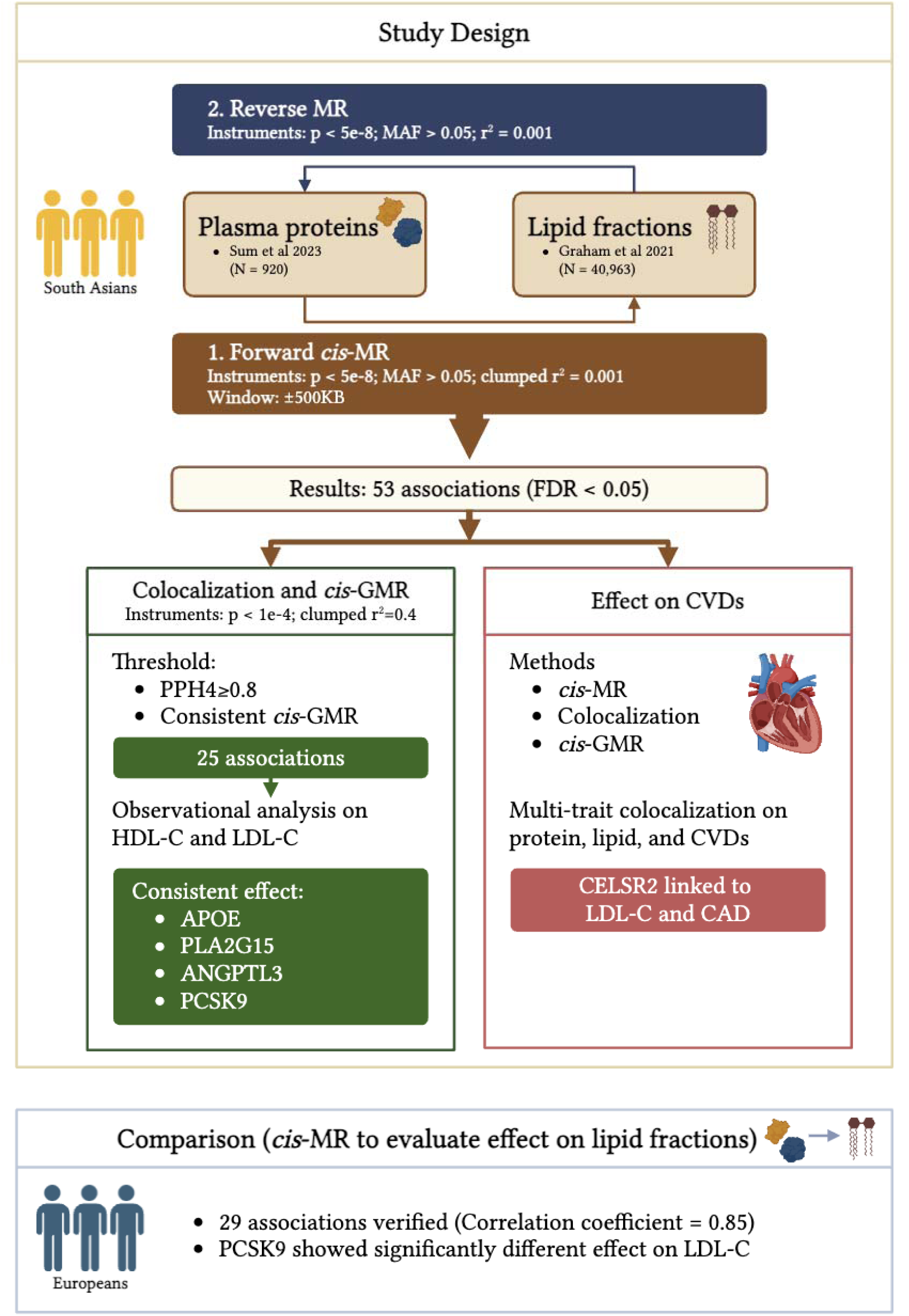
Overview of the study design. MR; Mendelian randomization, FDR; false discovery rate, GMR; generalized MR, PPH4; posterior probability of hypothesis 4, HDL-C and LDL-C; high- and low-density lipoprotein cholesterol

### Proteome-wide MR identified 29 plasma proteins associated with lipid fractions in SAS

Excluding proteins with ambiguous cognate genes and those encoded by genes located on the X-chromosome or within the *MHC* region (**Table S1**), 2,800 proteins were included in our study. Of these, 708 had at least one genetic instrument variable available (**Figure S1**) and were carried forward for the proteome-wide MR study. Using the Wald ratio (IV=1) or IVW (IVs≥2) approach as the primary MR method, 186 plasma proteins showed a potential causal effect on at least one of the lipid fractions (309 associations in total; P < 0.05; **Table S2**). After adjusting for multiple testing, a total of 29 genetically proxied plasma proteins showed potential causal effect on at least one of the 5 lipid fractions (53 associations in total; FDR < 0.05, **Figure 2, Table S2**).

**Figure 2.**
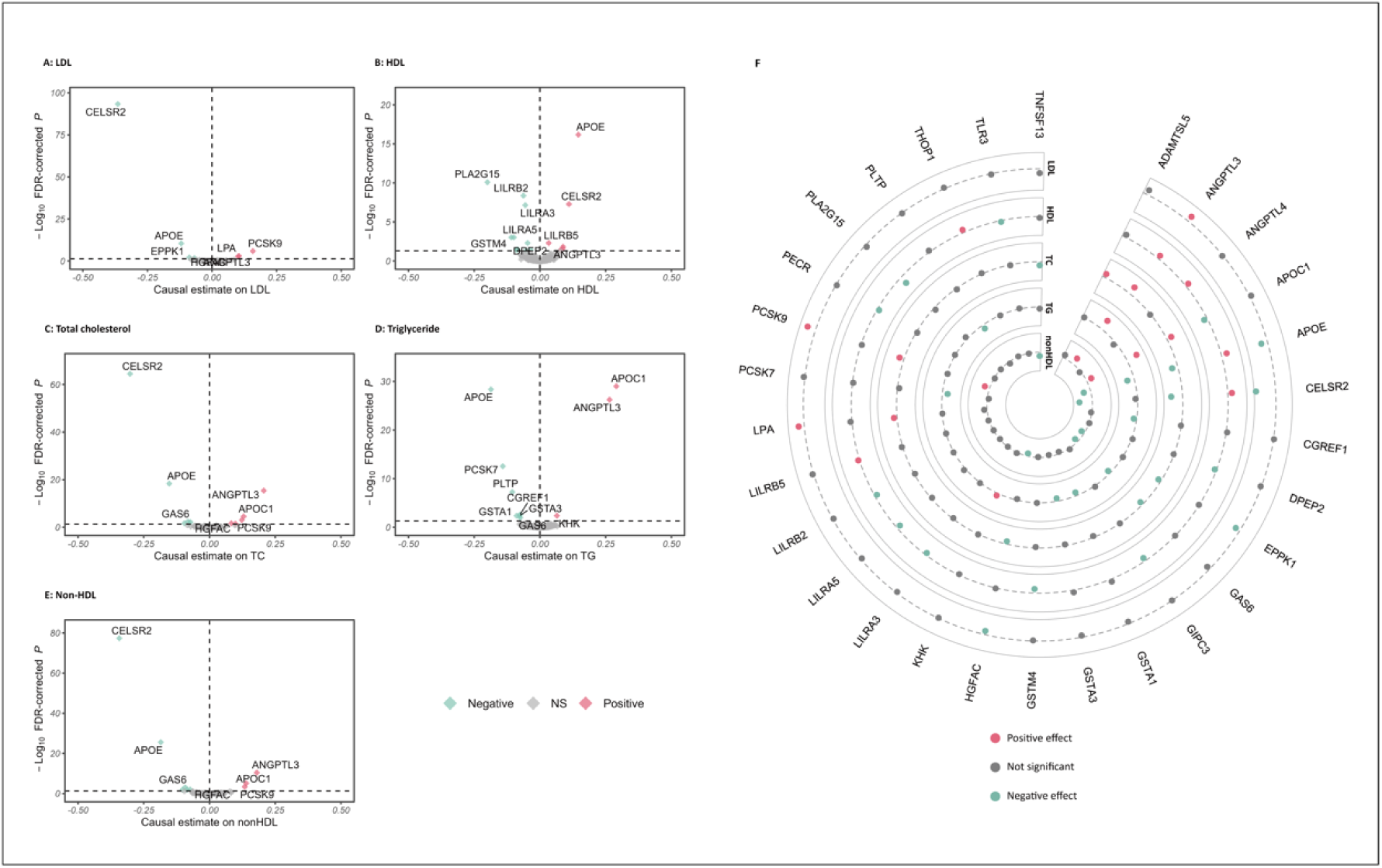
Volcano plots showing the causal effect of circulating plasma protein on the 5 lipid traits A) LDL-C; B) HDL-C; C) TC; D) TG; E) non-HDL-C. Each dot indicates a plasma protein with the x-axis showing the Wald ratio or IVW while the y-axis showing -log_10_ FDR-corrected P from the MR analysis. F) Circular plot showing the overlap of plasma proteins with the 5 lipid traits tested.

To avoid bias due to sample overlap between UKBPPP and GLGC, we performed MR using GLGC data without UKBB participants and the MR estimates were largely consistent across GLGC with and without UKBB (Pearson r^2^ = 0.93, P< 0.001; **Figure S2A, Figure S2B**).

Among these 29 lipids-associated proteins (53 associations), 12 (25 associations) were supported by strong colocalization evidence (PPH4 > 80%) with one of the lipids fractions including ANGPTL3, APOE, CELSR2, EPPK1, GAS6, GSTA1, GSTA3, HGFAC, LPA, PCSK9, PLA2G15, and PLTP (**Table S3**). Particularly, we identified strong colocalization of ANGPTL3 with all lipid fractions except HDL, CELSR2 with all lipid fractions except TG, LPA with LDL-C and TC, and PCSK9 with HDL, TC, and non-HDL. Additionally, 7 plasma proteins (7 associations in total) showed suggestive colocalization evidence (80% > PPH4 ≥ 60%) with the tested lipid fractions (**Figure 2**, **Figure S3, Table S3**).

To further validate our findings in the proteome-wide cis-MR, we included moderately correlated SNPs (r^2^ < 0.4) that are adequately associated with the plasma proteins (p < 1 × 10^-4^) and applied gIVW (36). Out of the 53 associations of lipid fractions with plasma proteins, gIVW produced consistent estimates with FDR corrected *P* value < 0.05 for 43 associations. Subsequently, gEgger and Weighted median were applied to the 42 associations with≥ 3 SNP instruments. Weighted median produced highly consistent estimates for all 42 associations. The gEgger detected no horizontal pleiotropy (FDR corrected *P* for intercept < 0.05) but derived inconsistent estimates for the association of TNFSF13 with TC and nonHDL-C, THOP1 with HDL-C, and EPPK1 with TC (**Figure 2**, **Table S4**).

Altogether, 30 associations of 14 genetically proxied proteins with lipid fractions were identified by proteome-wide MR, further validated by colocalization (either strong or suggestive) and the subsequent MR analysis generalized to correlated instruments (GMR). The top findings include CELSR2 associated with all lipid fractions except TG, PCSK9 and HGFAC with TC, LDL-C, and non-HDL-C, LPA with LDL-C and TC, and ANGPTL3 with all lipid fractions (**Figure 2**).

### Potential causal effects of lipid-associated proteins on CVD outcomes

Subsequently, we investigated whether lipid-associated proteins (N=29) identified by proteome-wide MR have potential causal effects on risk of CAD and stroke in SAS. After correcting for multiple testing, only genetically predicted CELSR2 had a causal association with CAD (Odds ratio (OR) = 0.64, 95% Confidence Interval (CI) = (0.50, 0.81), FDR = 0.003; **Figure 3A, Table S5**) which was also supported by strong colocalization evidence (PPH4 = 93.6%, **Figure 3B**, **Table S6**). Additionally, gIVW, gEgger, and weighted median produced consistent estimates and no pleiotropy was detected (**Figure 3C**, **Table S7**). Notably, ANGPTL3 and LPA showed suggestive associations with CVDs (P< 0.05) but did not pass the 5% FDR threshold (**Figure 3A**).

**Figure 3.**
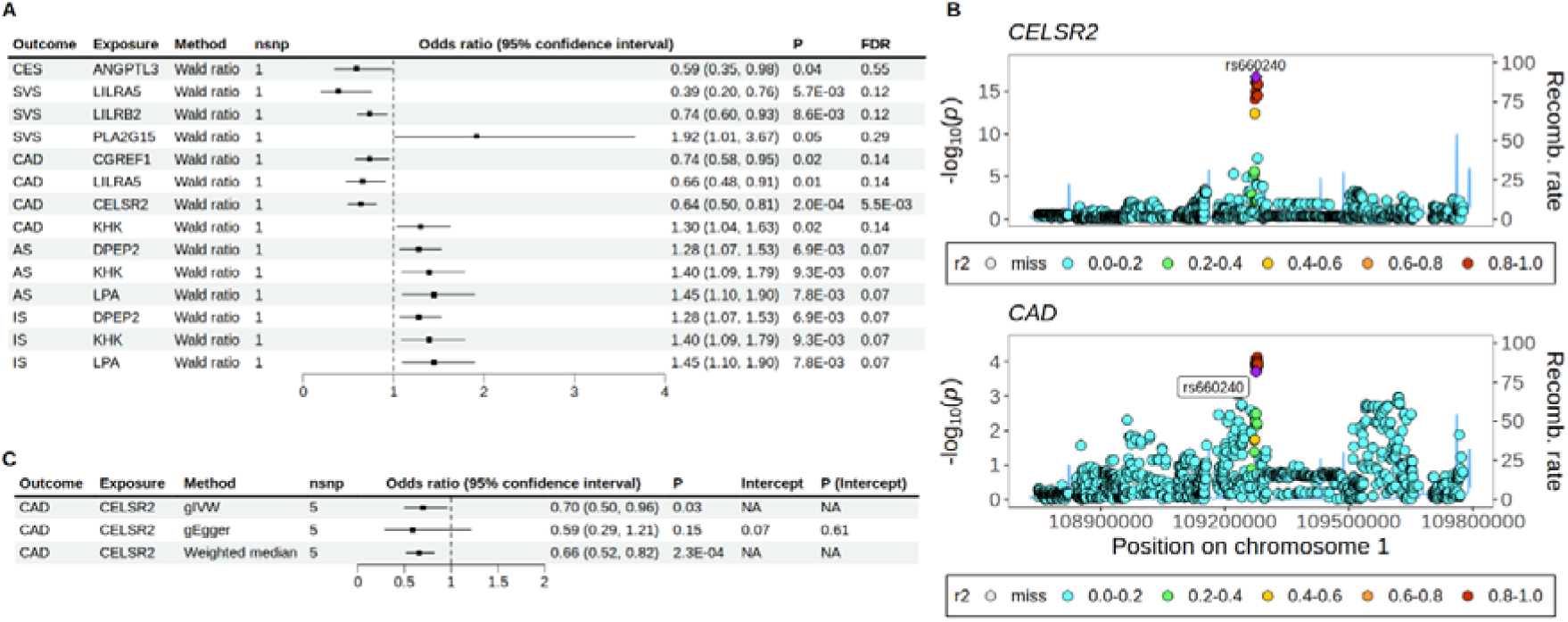
Effect of lipid-associated plasma proteins on CVDs A) Forest plot showing all plasma proteins that were nominally associated with CVDs (P< 0.05). B) Stacked genomic plot showing evidence for colocalization between CELSR2 and CAD; and C) Forest plot showing the effect of CELSR2 on CAD estimated by gIVW, gEgger, and weighted median.

Since the efficacy of lipid-associated proteins may be prioritized depending on the association of lipid fractions with CVDs, we assessed the causal effects of five lipid fractions on CVDs to indicate proteins more promising to CVD treatment. Our univariable MR identified four associations reaching nominal significance (P < 0.05), including LDL-C with CAD (β_IVW_ = 1.64; 95% CI = (1.03, 2.62) and cardioembolic stroke (β_IVW_ = 1.78; 95% CI = (1.01, 3.15); **Table S8**). The MR-Egger and weighted median methods produced estimates consistent in direction with the inverse variance weighted method (P< 0.05).

Since CELSR2 showed potential causal effects on both LDL-C and CAD while genetically proxied LDL-C was also causally associated with CAD risk, a multi-trait colocalization was performed on the 3 traits in the genomic region +/- 500KB extended from the CELSR2 gene. The multi-trait colocalization produced a posterior probability of 70.0% that CELSR2, LDL- C, and CAD colocalized in this region (**Figure 4**, **Table S9**). The posterior probabilities for all 15 scenarios of multi-trait colocalization were presented in **Table S9**.

**Figure 4.**
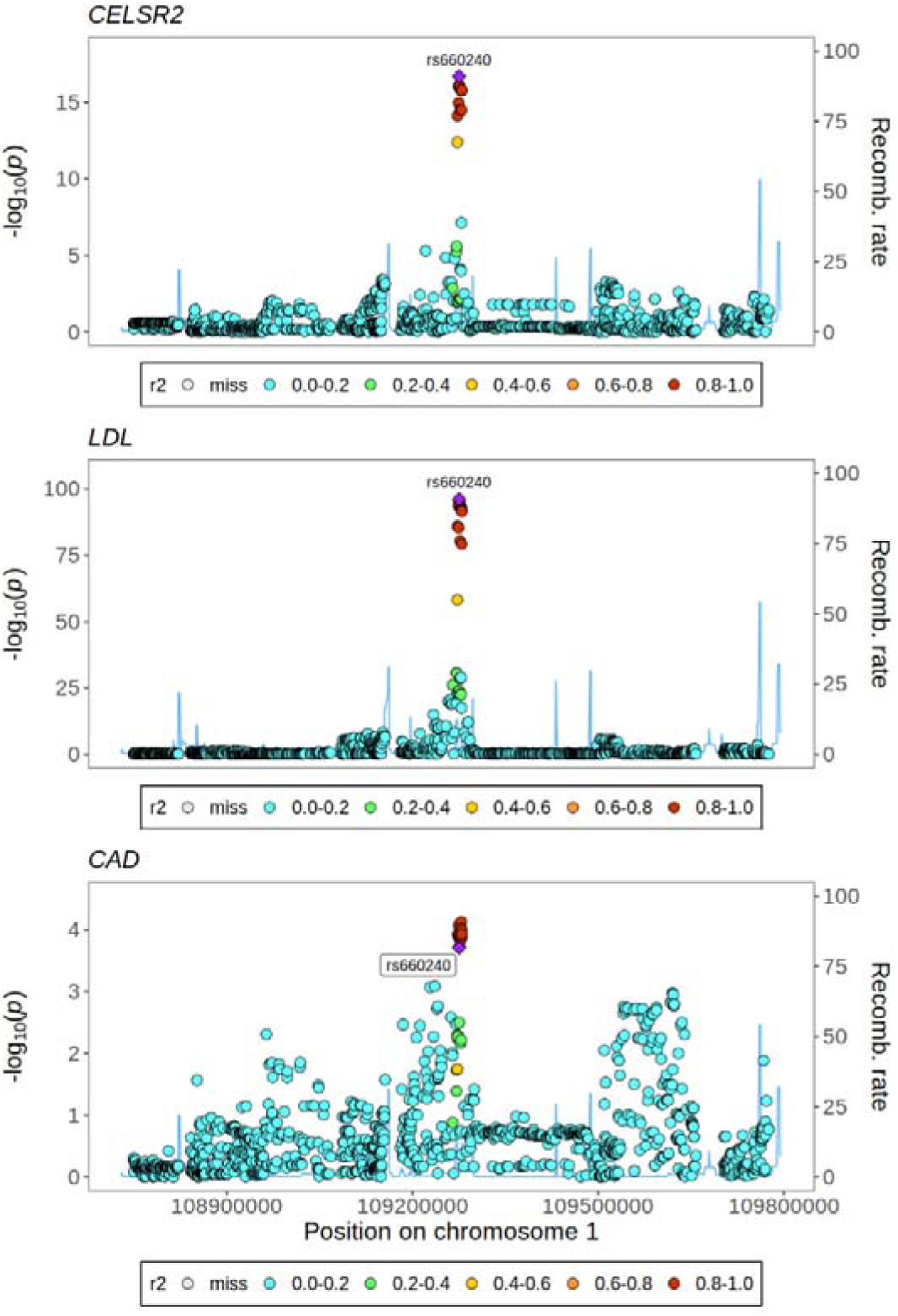
Stacked regional genomic plot from multi-trait colocalization showing the colocalized genetic variant rs660240 across LDL-C, CAD at the CELSR2 locus in SAS.

### Reverse MR with lipid fractions as exposures and plasma proteins as outcomes

To understand the plasma proteins modified by lipid fractions, a reverse MR was performed using lipid fractions as exposures and plasma proteins as outcomes. This analysis identified genetically proxied associations between TG and LDLR, and HDL-C with 3 proteins: APOA1, MENT, and FGFBP2 (β_range_ = 0.55 to 0.61, FDR-corrected P< 0.05**, Table S10, Figure S4**). Subsequent MR-Egger and weighted median produced consistent estimates and detected no horizontal pleiotropy (**Table S11, Figure S5**).

### Comparison with EUR population

Among the 53 proteome-wide MR identified associations in SAS, 29 of them had consistent beta estimates in EUR (FDR-corrected P < 0.05). The correlation analysis on the 30 beta estimates in SAS against those in EUR produced a correlation coefficient of 0.85 (*P* = 6.5 ×10 ^-9^, **Figure S6).** Out of the 30 associations in SAS with MR, colocalization, and GMR evidence, 22 were verified in EUR population (**Table S12** and **Figure 5**). Proteome-wide MR, colocalization, and GMR identified 6 proteins associated with HDL-C in SAS, but only ANGPTL3 and APOE were also significant in EUR (**Figure 5**). Of the 6 proteins linked to LDL-C in SAS, ANGPTL3, CELSR2, LPA, and PCSK9 were verified in EUR, with PCSK9 showing a stronger effect in EUR (β_EUR_ = 0.37; 95% CI = (0.36, 0.38), β_SAS_ = 0.16; 95% CI = (0.11, 0.21); **Figure 5**). Among the remaining 18 associations with nonHDL-C, TC, or TG, only the association of EPPK1 with nonHDL-C and HGFAC with TC were not significant in EUR.

**Figure 5.**
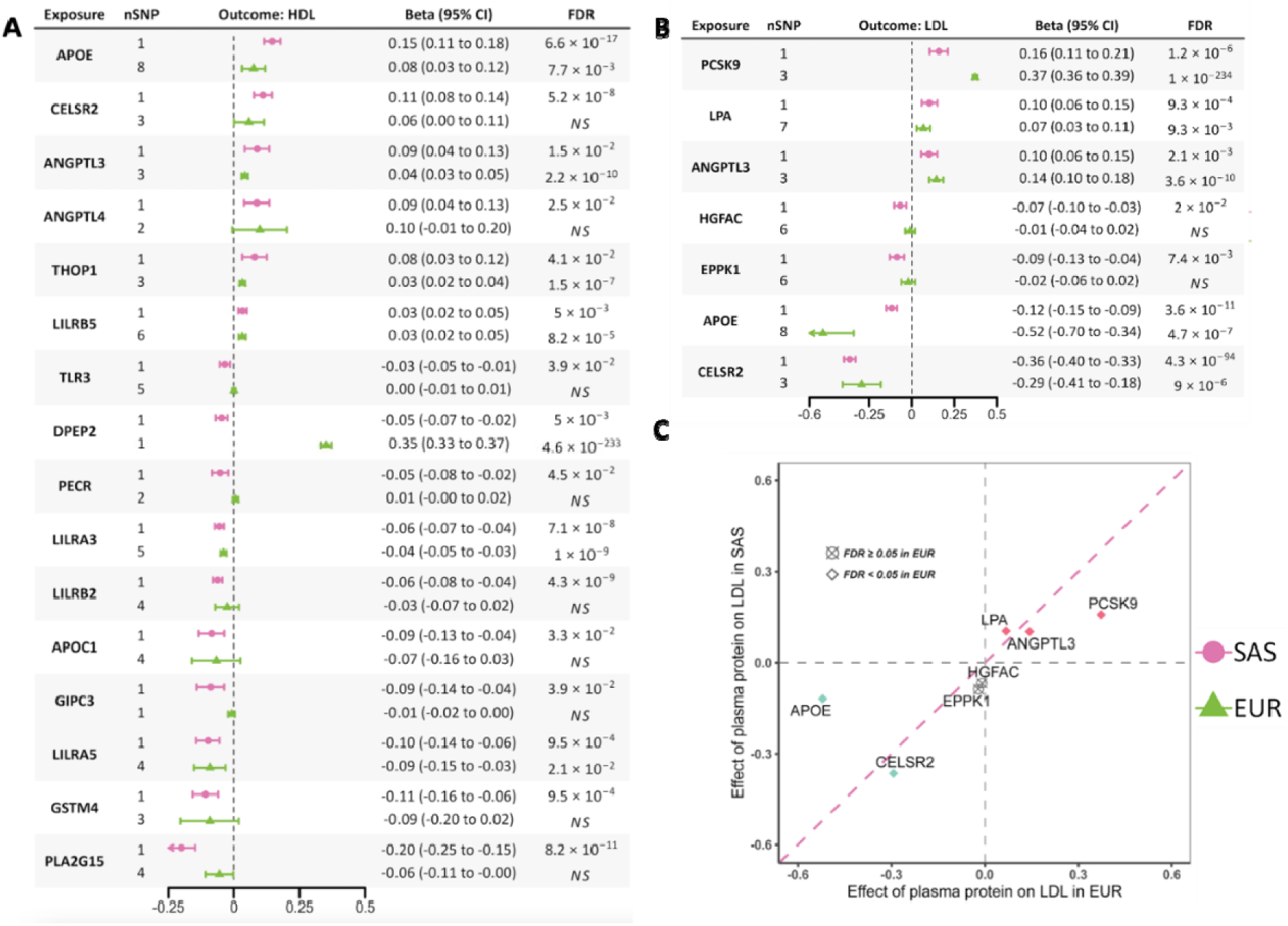
Effect of plasma proteins on lipid fractions using cis-MR in SAS and EUR on A) HDL-C; B) LDL-C; and C) Scatter plot for comparison of causal effect estimates from MR between EUR and SAS for LDL-C.

### Observational associations of protein levels with lipid fractions

There were 3 proteins with strong evidence (MR, strong colocalization, and GMR) of association HDL-C (APOE, CELSR2, and PLA2G15) and 6 proteins with LDL-C (ANGPTL3, CELSR2, EPPK1, HGFAC, LPA, and PCSK9) that were tested in observational analysis. After adjusting for multiple testing, all 9 proteins had a significant association with their genetically associated lipid fraction (FDR adjusted *P* < 0.05) but only 4 had a consistent direction of effect with MR estimates **(Table S13)**; APOE (β = 0.078; 95% CI = (0.075, 0.082)) and PLA2G15 (β = -0.067; 95% CI = (-0.071, -0.063)) with HDL, and ANGPTL3 (β = 0.180; 95% CI = (0.175, 0.184)) and PCSK9 (β = 0.196; 95% CI = (0.192, 0.200)) with LDL. Out of these four proteins, there was a significant interaction between ANGPTL3 and PCSK9 levels with LDL-Cin South Asians with South Asians having a significantly lower effect compared to other ancestries (ANGPLT3*South Asian; β = -0.072; 95% CI = (-0.103, - 0.040), PCSK9*South Asian; β = -0.140; 95% CI = (-0.174, -0.107)).

## Discussion

### Key findings in this study

Here, we performed a bidirectional proteome-wide MR on five lipid fractions, and linked lipid-related proteins to cardiovascular outcomes in SAS. Our study confirmed key proteins (PCSK9, ANGPTL3, LPA), identified novel targets (GSTA1, GSTA3, EPPK1, PECR, and PLA2G12), and strengthened evidence for CELSR2 and GAS6 in dyslipidaemia. Notably, our results highlight significant heterogeneity in MR estimates across genetic ancestry groups, particularly for the effect of PCSK9 on LDL-C. We also report CELSR2 with evidence for its effect on LDL-C and CAD risk. Reverse MR identified LDLR as modifiable by TG, and APOA1, MENT, and FGFBP2 by HDL-C.

### Enhanced role of CELSR2 and GAS6 in lipid metabolism and cardiovascular outcomes

We found an inverse association of genetically proxied CELSR2 with LDL-C and CAD risk in SAS. CELSR2 is a transmembrane protein belonging to the flamingo family of cadherin superfamily (40). Although the biological function of CELSR2 is not well understood, the role of CELSR2 in lipid metabolism was indicated by some previous studies. A locus in the vicinity of CELSR2, rs599839 (in LD with rs660240, the instrument of CELSR2 in this study (r^2^ = 0.989 in SAS, r^2^ = 0.871 in EUR)), was first reportedly associated with CAD, LDL-C and TC by 2 European ancestral GWAS (41–43). rs660240, a 3’ UTR variant, is an eQTL for *CELSR2*, *PSRC1*, and *SORT1* in liver tissue (Open Target Genetics). It shows slight variations in allele frequencies between South Asians, Europeans, and East Asians, which could have implications for studies related to disease susceptibility and treatment response. Furthermore, a transcriptomic study revealed the risk allele of rs599839 to CAD and high LDL-C also suppressed the expression of *CELSR2* gene in liver (44). Extending to non- European populations, the association of *CELSR2* variants with lipid fractions and CAD risk was also verified in the South Asian population (45, 46). However, although the effect of CELRS2 on lipid metabolism was indicated by genetic and transcriptomic studies, the underlying mechanism is less clear. One study demonstrated that CELSR2 deficiency can elevate reactive oxygen species of hepatocytes, which impairs lipid homeostasis and physiological unfolded protein response(47). In conclusion, our result is consistent with the significant role of CELSR2 in CAD and lipid metabolism suggested in earlier studies. Our study also extends the generalizability of CELSR2’s function to the South Asian population.

We identified an inverse association of genetically proxied GAS6 with TC and TG. GAS6 is a ligand for TAM receptor protein tyrosine kinases including AXL, TYRO3 and MER. The GAS6 - TAM pathway was found implicated in carcinoma, inflammation, and haemostasis and has been targeted for the treatment of carcinoma (48–50). In recent years, there is also evidence that GAS6 plays a role in regulating obesity and lipid metabolism (51, 52) as plasma gamma-glutamyl carboxylated GAS6 (Gla-GAS6) was found significantly lower in hyperlipidaemic individuals compared with healthy controls (51). The subsequent experiment showed that higher Gla-GAS6 expression induced by vitamin K in plasma and hepatocyte could reduce the plasma lipid level in hyperlipidaemic mice (51). The Gla-GAS6 takes effect by regulating the AMPK/SREBP1/PPARα signalling pathways of hepatic lipid metabolism (51). Our study supports this hypothesis by providing genetic evidence for GAS6 as a potential regulator of lipid metabolism.

### Known targets and novel findings in lipid metabolism

In this study, we replicated previously established protein associations with lipid metabolism, including ANGPTL3, APOE, LPA, PCSK9, and PLTP, some of which are drug targets for dyslipidemia treatment (53). However, despite the concordance in direction and significance for the association of PCSK9 on LDL-C in both SAS and EUR, we identified a significantly reduced effect in SAS as supported by both MR and observational analysis. Since drug responses to lipid-lowering therapy can vary across ethnicities, this finding may have important clinical implications. Previous studies suggest that atorvastatin and simvastatin have similar lipid-lowering effects in SAS patients compared to those in EUR (54). Therefore, PCSK9, as a novel target for lipid-lowering medication, warrants further investigation for their effect in more diverse ethnicity. Mechanisms accounting for the ancestral heterogeneity in PCSK9’s effect are limited. However, a recent study sequencing PCSK9 gene in Indians indicated difference in prevalence of mutation in SAS. The by-ancestry heterogeneous mutation pattern can result in heterogeneity of PCSK9 structure and activity, which may modify the effect of PCSK9 abundance (55).

Additionally, our study identified novel associations, including HGFAC, GIPC3, EPPK1, GSTA1, GSTA3, PECR, and PLA2G12. HGFAC activates hepatocyte growth factor (HGF) by converting it to a heterodimer, which then binds to the MET receptor to activate downstream signalling. A previous study linked a putative HGFAC loss-of-function variant to elevated serum TG and LDL-C (56). In another animal experiment, an increase of circulating TG was present in both male and female HGFAC-KO mice while higher level of circulating TC was present in male HGFAC-KO mice (57). The association of GIPC3 with HDL-C was firstly identified in our study. However, a previous study reported that GIPC1, another subtype of GIPC family, is involved in lipid metabolism by regulating the SR-B1 expression (58). Lastly, we also identified GSTA1, GSTA3, EPPK1, PECR, and PLA2G12 associated with lipid traits, all of which are novel findings.

### Reverse effect of lipid fractions on plasma proteins

Conversely, we applied MR to identify plasma proteins modified by lipid fractions. We found that genetically proxied TG levels were associated with increased plasma LDLR. LDLR is a cell membrane glycoprotein that regulates lipid homeostasis by binding and internalizing circulating cholesterol-containing lipoprotein particles, including LDL-C, VLDL-C, and chylomicron remnants (59). Deficiency in LDLR can result in dyslipidaemia. Therefore, we interpreted that the LDLR upregulation triggered by genetically proxied TG is likely to reverse the hyperlipidaemia. In addition, we observed genetically proxied HDL-C level associated with increased plasma APOA1, MENT, and FGFBP2 level. All 3 proteins play important roles in various disorders and mechanisms, including cholesterol transport, angiogenesis, tissue repair, and cellular metabolism. Therefore, further investigation is necessary for detailed biological interpretation of these findings.

### Strength and Limitations

Our study has several strengths. First, to the best of our knowledge, this is the first MR study to systematically evaluate the potential causal association between plasma proteins and lipid traits in South Asians, replicating known protein-stroke associations and discovering novel targets. Secondly, we combined MR with colocalization, to reduce bias from LD and reverse causation — a potential limitation of conventional observational studies. Finally, by incorporating GWAS on CVDs, we linked lipid associated proteins to cardiovascular outcomes and identified CELSR2 as a promising target for both LDL-C and CAD.

There are also limitations in our study. First, the genetic associations for plasma proteins in SAS were based on a relatively small sample size, compared with current GWAS standards, so our findings should be interpreted cautiously, and larger GWAS are needed for validation. Second, despite using various MR methods and sensitivity analyses, unaccounted pleiotropy may still bias our results. Third, our analysis assumes the absence of SNP-SNP and SNPCenvironment interaction. Lastly, our findings may not be generalizable to ancestry groups other than the ones studied here.

## Conclusion

Our comprehensive study triangulated evidence from MR, colocalization, and observational analyses, highlighting several novel proteins associated with lipid fractions in South Asians. Notably, our analysis suggests that the causal effect of PCSK9 on LDL-C may be ancestry- specific. Future studies with larger sample sizes are needed to validate our findings, along with further mechanistic and clinical studies to confirm the role of PCSK9 on LDL-C in South Asians and Europeans.

## Data Availability

The UKBPPP data can be downloaded from http://ukb-ppp.gwas.eu. GLGC dataset can be downloaded from https://csg.sph.umich.edu/willer/public/glgc-lipids2021/results/ancestry_specific/. ELGH data can be downloaded from https://www.genesandhealth.org/research/scientific-data-downloads/gwas-data-genes-health-feb-2020-datafreeze. GWAS summary statistics for stroke and its subtypes for South Asians and Europeans can be obtained from the GWAS catalogue with accession GCST90104559-GCST90104563, and GCST90104539-GCST90104543, respectively.

https://csg.sph.umich.edu/willer/public/glgc-lipids2021/results/ancestry_specific/

https://www.genesandhealth.org/research/scientific-data-downloads/gwas-data-genes-health-feb-2020-datafreeze

https://www.ebi.ac.uk/gwas/

## Contributors

DM, IT and AD contributed to the conception and design of the study. SW and AS contributed to acquisition and statistical analysis of data. DM, IT, AD supervised the project. All authors contributed to the drafting and critical revision of the manuscript.

## Acknowledgments

The authors would like to thank the staff and participants who contributed to the UK Biobank, UKBPPP, GLGC, the East London Genes & Health (ELGH) study, the CARDIoGRAMplusC4D consortium, GIGASTROKE consortium for making the resources available.

## Funding

This work is supported by the British Heart Foundation Research Excellence Award (4) (RE/24/130023). IT and AR are supported by NIH R01 HL162300C02.

## Conflict of interest

A.R. reported consultancies for Raydel, Johnson & Johnson, and JP Morgan, all with modest compensation, and received a significant research grant from CSL Limited but with no salary support included. The remaining authors have no disclosures to report.

