## Supplementary figures and images for "Prioritizing protein targets for dyslipidaemia and cardiovascular diseases using Mendelian randomization in South Asians"

### SF1_MR_IV_Count.tiff

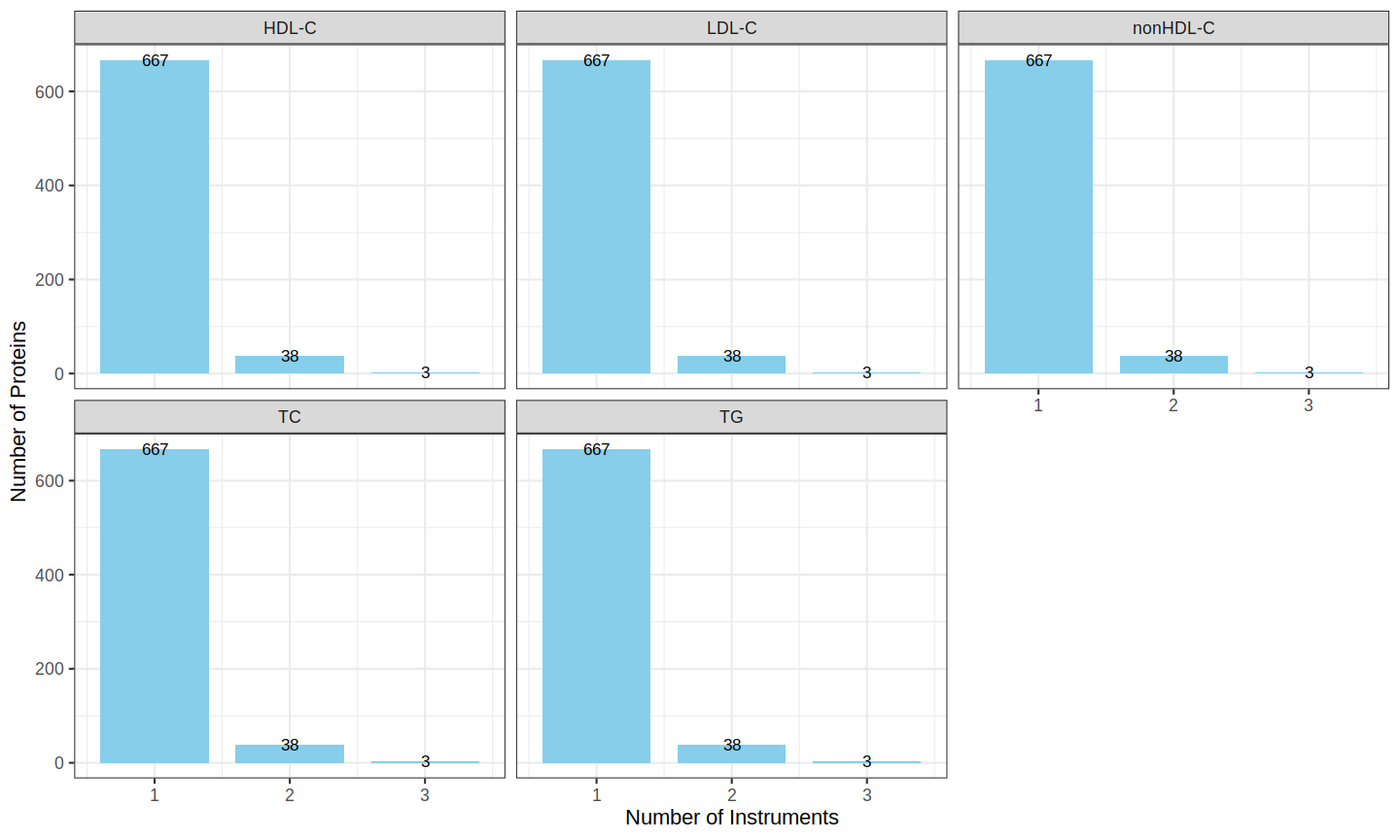

### SF2A_Scatterplot_beta_noUKB_vs_wzUKB.tiff

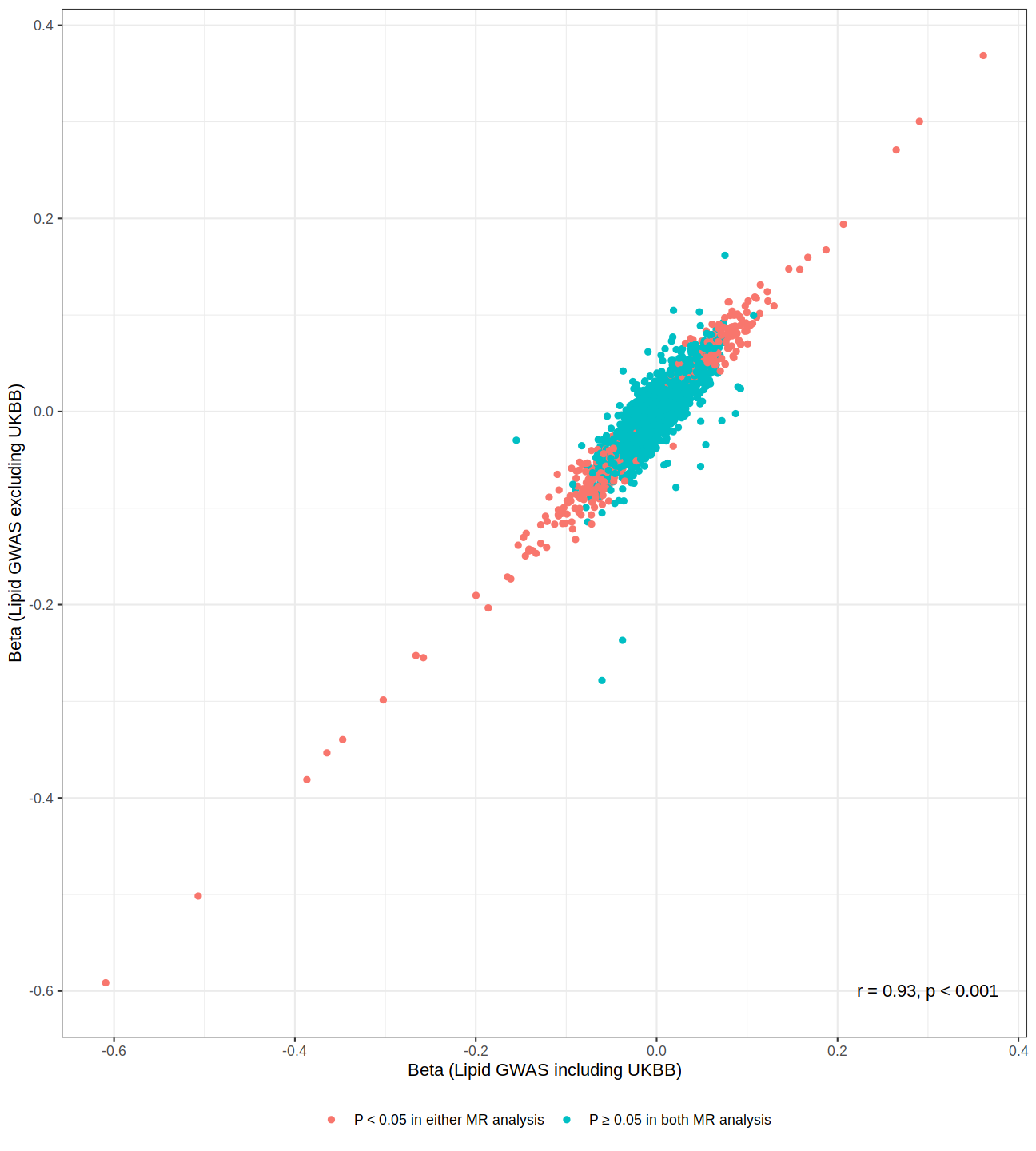

### SF2B_volcano_plot_noUKB_vs_wzUKB.tiff

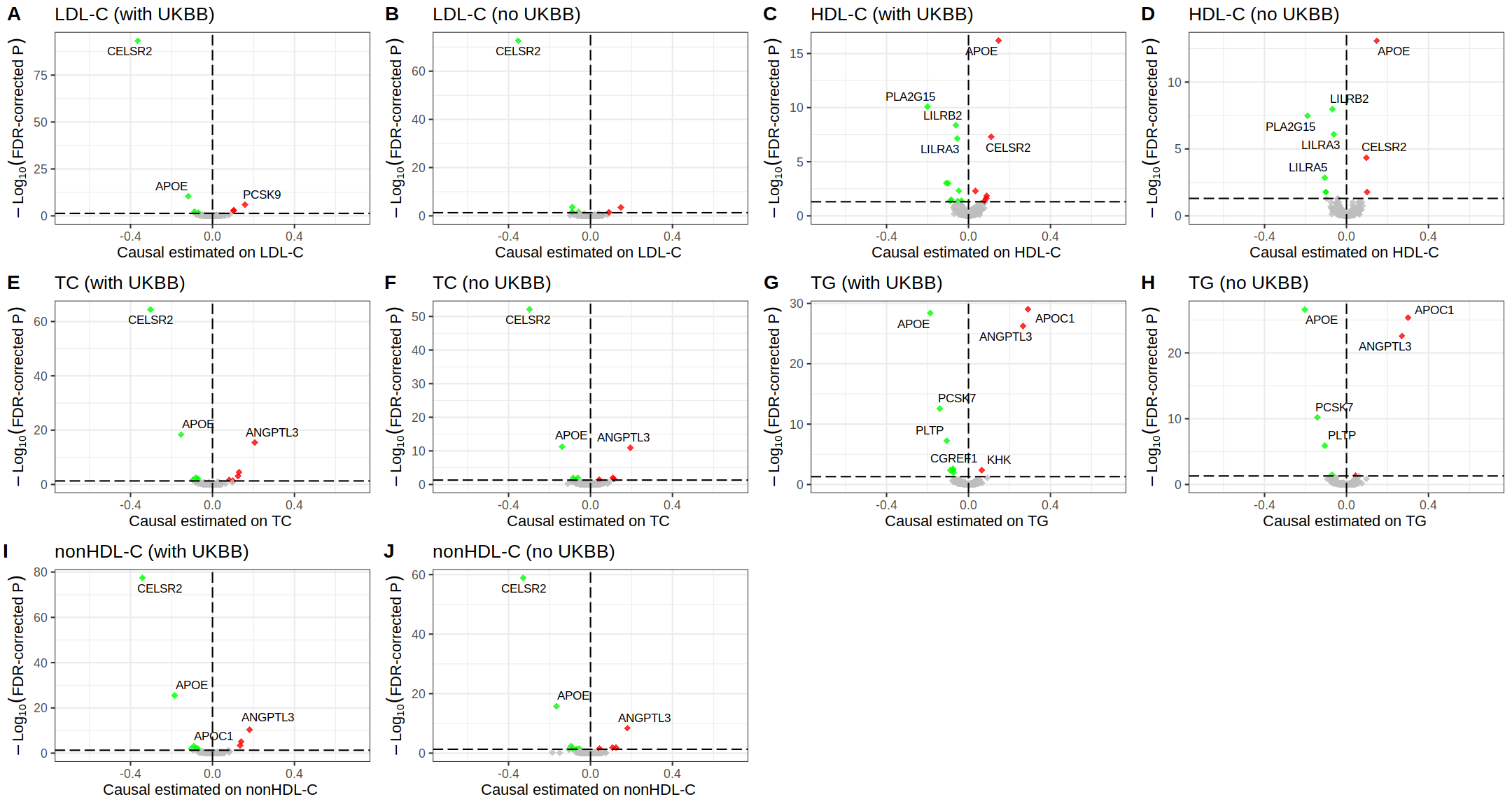

### SF3A_UKBPPP_SAS_LDL_Coloc_Stackedplot.tiff

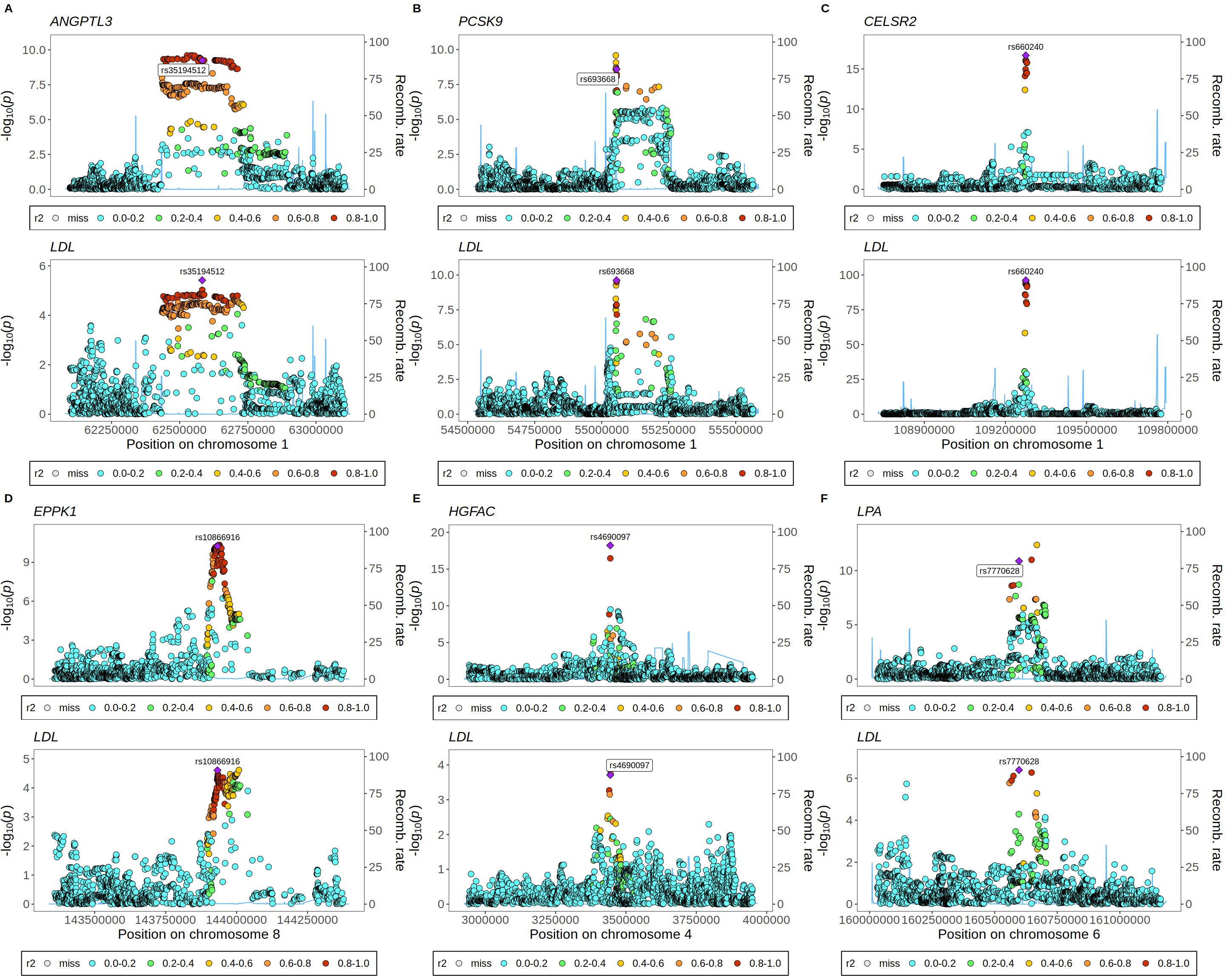

### SF3B_UKBPPP_SAS_HDL_Coloc_Stackedplot.tiff

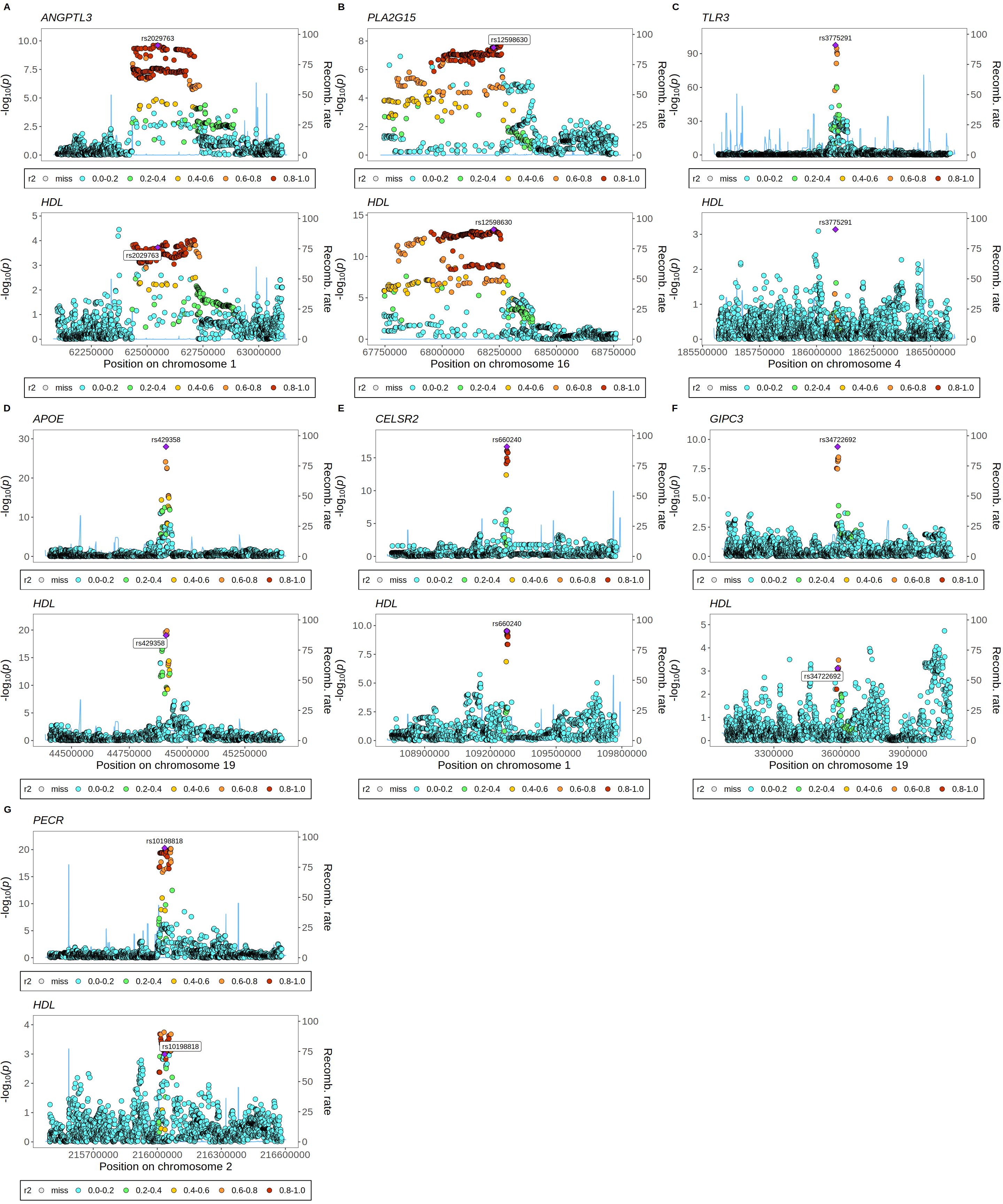

### SF3C_UKBPPP_SAS_TC_Coloc_Stackedplot.tiff

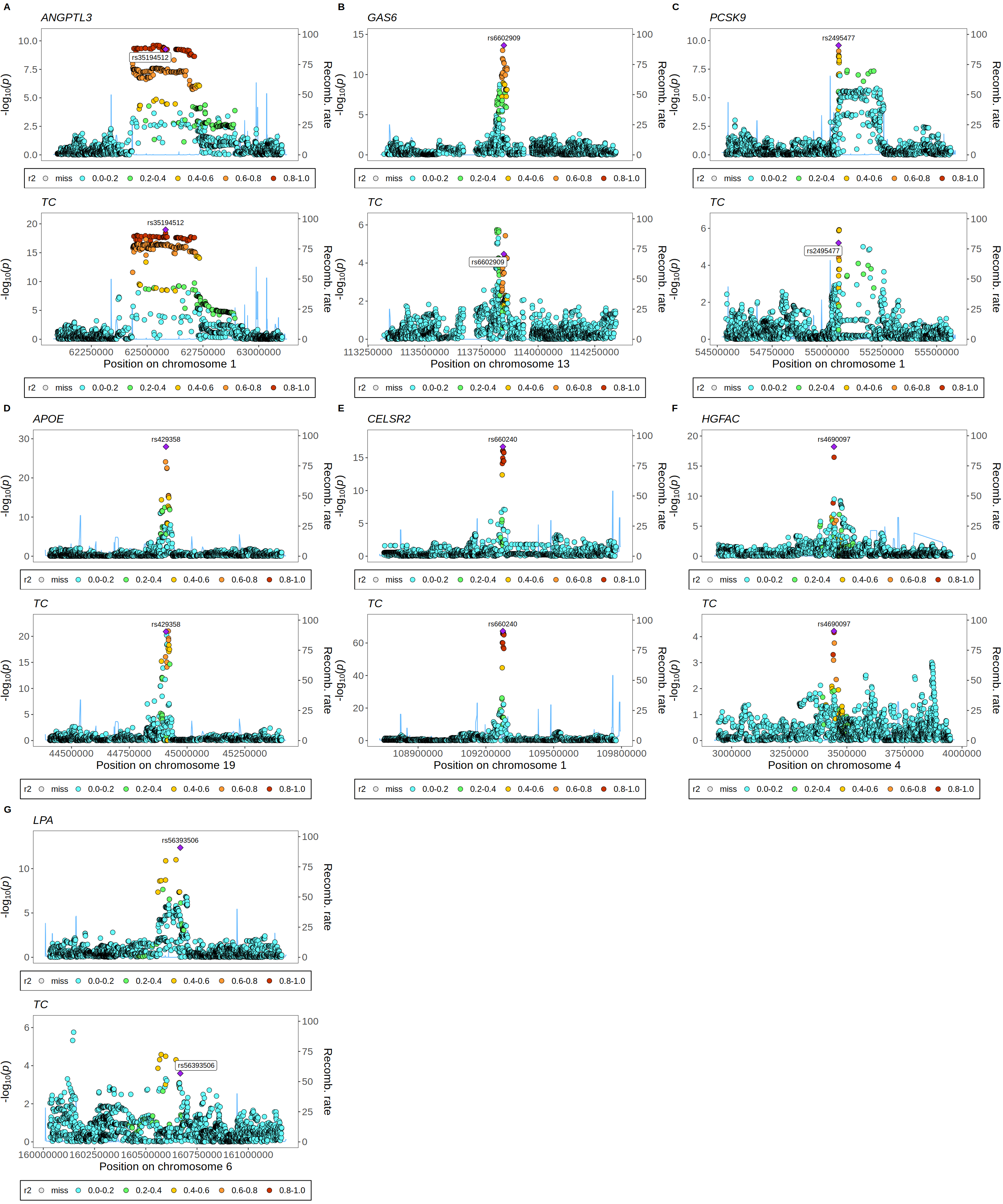

### SF3D_UKBPPP_SAS_TG_Coloc_Stackedplot.tiff

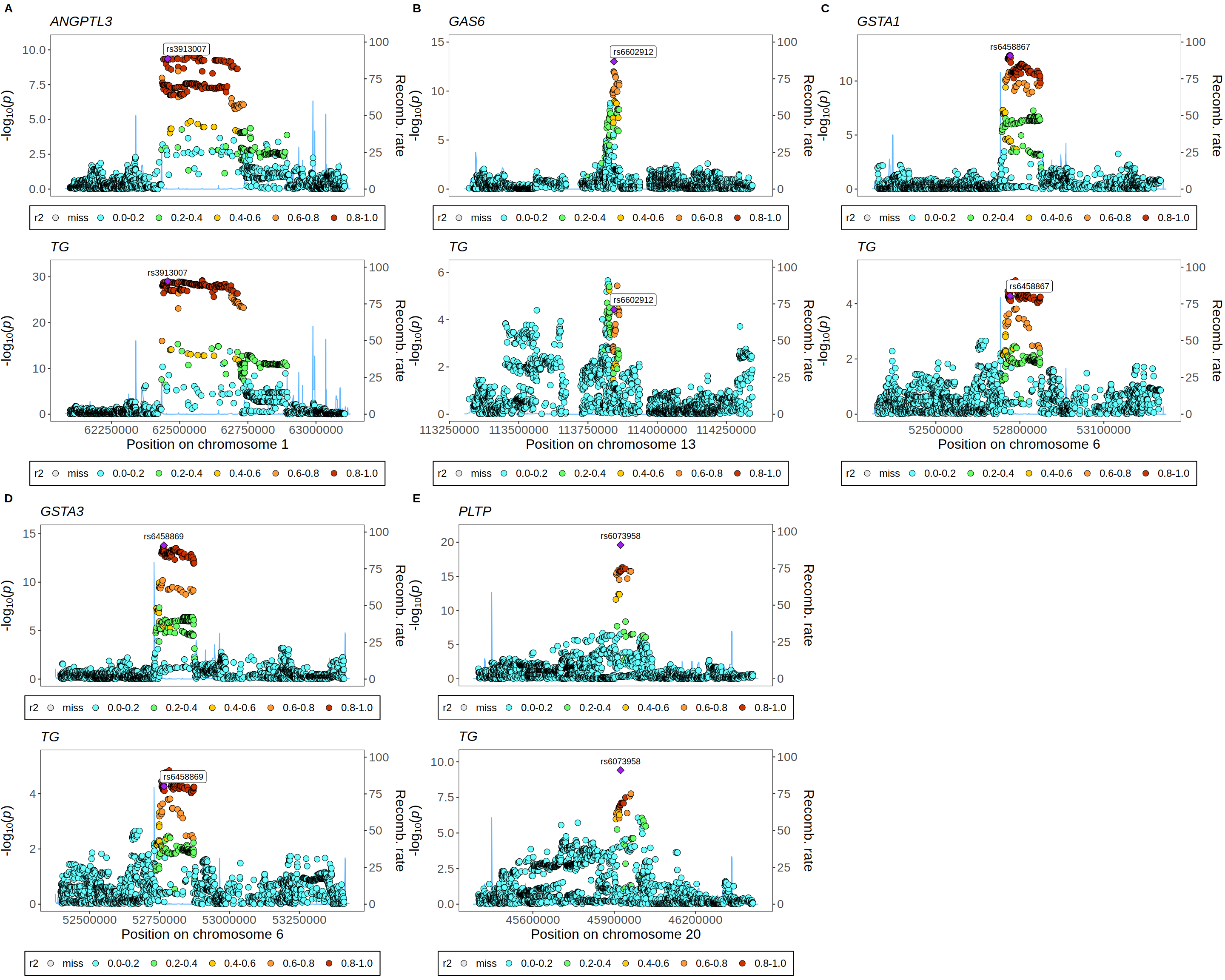

### SF4_Lipids_SAS_UKBPPP_MR.tiff

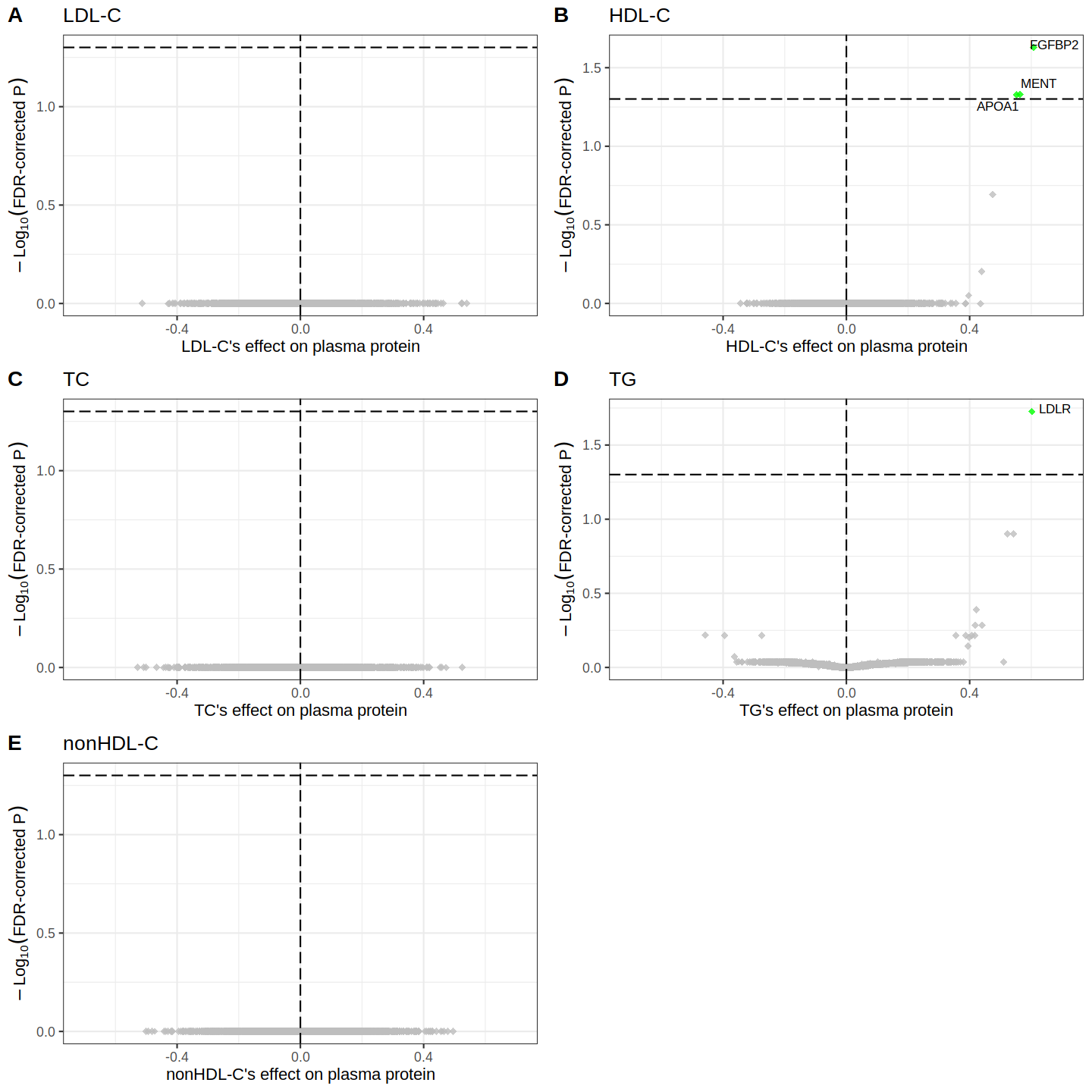

### SF5_Lipids_SAS_UKBPPP_RobustMR.tiff

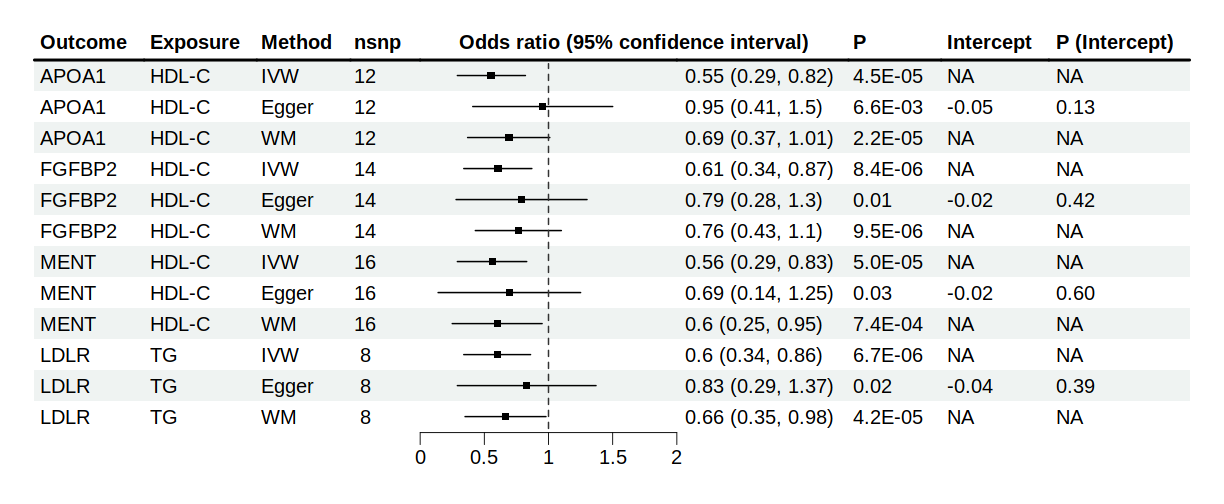

### SF6_Scatterplot_All_beta_EUR_vs_SAS.tiff

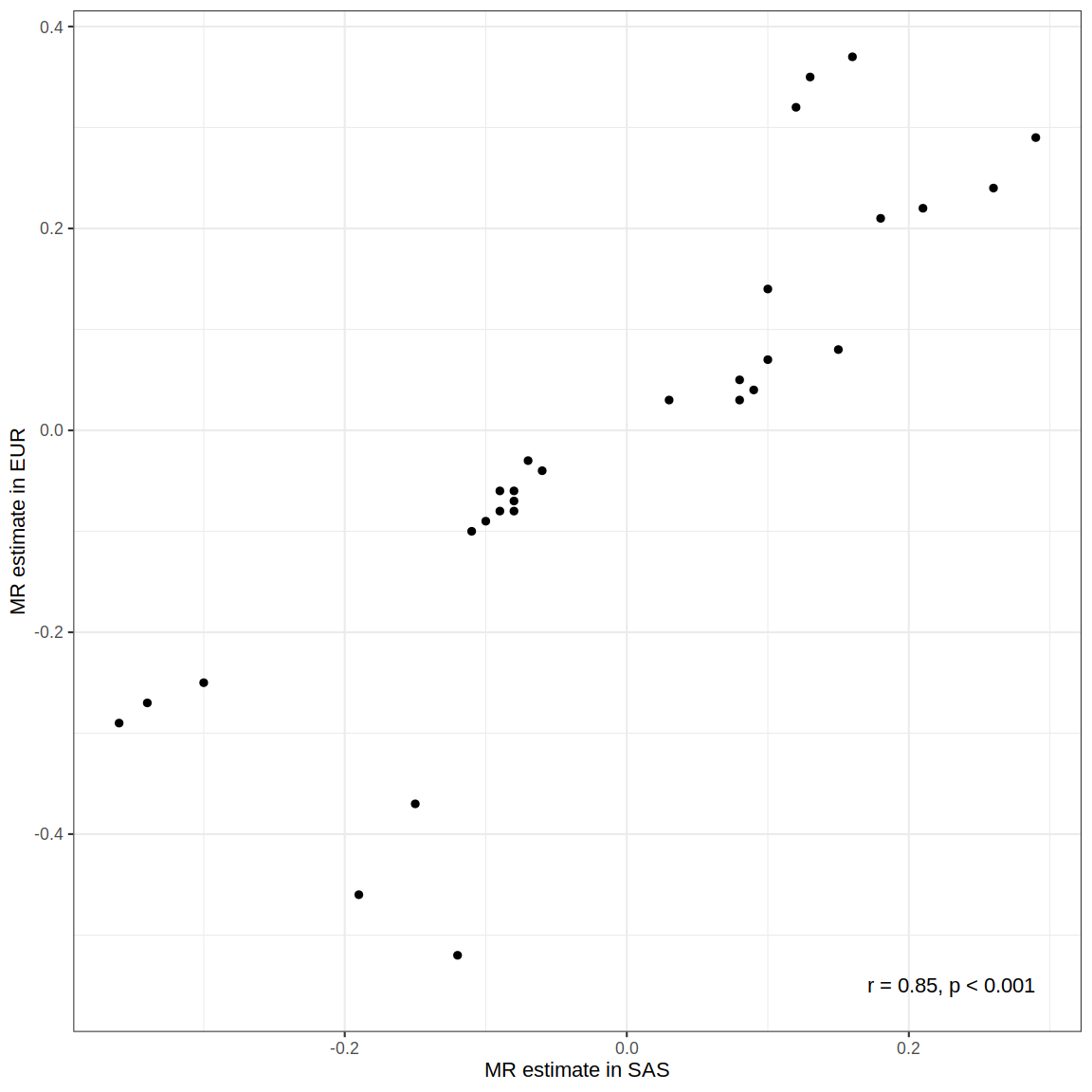
